# The U-shaped association of serum iron level with disease severity in adult hospitalized patients with COVID-19

**DOI:** 10.1101/2021.02.19.21252061

**Authors:** Kentaro Tojo, Yoh Sugawara, Yasufumi Oi, Fumihiro Ogawa, Takuma Higurashi, Yukihiro Yoshimura, Nobuyuki Miyata, Hajime Hayami, Yoshikazu Yamaguchi, Yoko Ishikawa, Ichiro Takeuchi, Natsuo Tachikawa, Takahisa Goto

## Abstract

Coronavirus disease 2019 (COVID-19) is an emerging infectious disease that leads to severe respiratory failure (RF). It is known that host exposure to viral infection triggers an iron-lowering response to mitigate pathogenic load and tissue damage. However, the association between host iron-lowering response and COVID-19 severity is not clear. This two-center observational study of 136 adult hospitalized COVID-19 patients analyzed the association between disease severity and initial serum iron, total iron-binding capacity (TIBC), and transferrin saturation (TSAT) levels. Serum iron levels were significantly lower in patients with mild RF than in the non-RF group; however, there were no significant differences in iron levels between the non-RF and severe RF groups, depicting a U-shaped association between serum iron levels and disease severity. TIBC levels decreased significantly with increasing severity; consequently, TSAT was significantly higher in patients with severe RF than in other patients. Multivariate analysis including only patients with RF adjusted for age and sex demonstrated that higher serum iron and TSAT levels were independently associated with the development of severe RF, indicating that inadequate response to lower serum iron might be an exacerbating factor for COVID-19.

## Introduction

Coronavirus disease 2019 (COVID-19) is an emerging infectious disease caused by severe acute respiratory syndrome coronavirus 2 (SARS-CoV-2), provoking worldwide pandemic emergencies. The main organ system affected by SARS-CoV-2 infection is the lungs, and in severe cases, acute respiratory distress syndrome (ARDS) with life-threatening respiratory failure occurs. Although the pathophysiology of COVID-19 has not been fully elucidated, complex interactions between the virus and host responses seem to account for disease severity^1^.

Host exposure to infection triggers multiple responses to mitigate pathogenic load and tissue damage, termed “resistance” and “tolerance,” respectively^2^. Immunological responses exerted by innate and acquired immune systems are among the best-known host resistance responses. Another host resistance strategy is the deprivation of molecules necessary for the pathogens to survive or replicate. Iron ions are among the nutrients necessary for viral replication^3,4^. When a host is infected with the virus, hepcidin is released from the liver and lowers serum iron levels to limit iron availability^3,5^. Moreover, iron overload induces oxidative stress and subsequent tissue damage^6^. Therefore, lowering iron levels also engages host tolerance responses to protect the host itself. In fact, it has been reported that high serum iron level is associated with worse mortality in patients with sepsis^7^.

Several previous reports have shown that a decreased serum iron level is observed in patients with severe COVID-19, and serum iron levels can be a prognostic marker^8–11^. However, the association between host iron-lowering response and disease severity is not clear. This study evaluated the association between initial iron metabolism indicators and disease severity in hospitalized COVID-19 patients with or without respiratory failure.

## Materials and Methods

### Study Design

In this two-center retrospective and prospective observational study, we analyzed the concentrations of total iron, total iron-binding capacity (TIBC), transferrin saturation (TSAT), and ferritin in the serum of patients with COVID-19. Data from patients admitted to Yokohama City University Hospital (YCUH) from April 1st to August 7th, 2020 and data from those admitted to Yokohama Municipal Citizen’s Hospital (YMCH) from April 1st to September 11th, 2020 were retrospectively collected. Whereas data from patients admitted to YCUH from August 8th, 2020 to January 26th, 2021 were prospectively collected. The study protocol was reviewed and approved by the institutional review boards of YCUH (Yokohama City University Certified Institutional Review Board, approval number: B200700099) and YMCH (Yokohama Municipal Citizen’s Hospital Institutional Review Board, approval number: 20-09-04). The need for informed consent was waived by Institutional Review Boards because of the observational design of the study. The study was conducted in accordance with the Declaration of Helsinki.

### Patients

The inclusion criteria were as follows: 1) aged ≥18 years, 2) COVID-19 diagnosis based on positive results of real-time polymerase chain reaction or SARS-CoV-2 antigen test, and 3) serum iron level measured within first 5 days after hospitalization. Patients who refused mechanical ventilation were excluded from the study.

### Outcome

Patient outcomes were classified into three categories according to their worst respiratory status during hospitalization as follows: non-respiratory failure (non-RF), not requiring oxygen therapy or mechanical ventilation (MV) throughout hospitalization; mild RF, requiring oxygen therapy or MV but with the worst arterial oxygen partial pressure / fractional inspired oxygen (P/F) ratio measured with MV or high-flow nasal oxygen therapy (HFNO) maintained above 200; severe RF, the worst P/F ratio measured with MV or HFNO ≤ 200.

### Statistical Analysis

All data are presented as medians ± interquartile ranges (IQR). We compared iron metabolism indicators among the three groups using the Kruskal-Wallis test followed by Dunn’s multiple comparison test. Clinical characteristics and laboratory values were analyzed with the Kruskal-Wallis test (continuous values) or the chi-square test (categorical values). Moreover, we analyzed the association between serum iron level or TSAT and disease severity among only patients with RF using a multivariate logistic regression model adjusting for patient age and sex. In the supplementary analysis, body mass index and the histories of hypertension, diabetes mellitus, and chronic respiratory diseases were also included as covariates in the multivariate logistic regression model. The statistical significance level was set at p < 0.05. All statistical analyses were performed using Prism software (version 9.0; GraphPad Software, San Diego, CA, USA).

## Results

### Patient Characteristics

One hundred thirty-six patients were included in the study among 204 patients with COVID-19 admitted to YCUH (59 patients) or YMCH (77 patients) (Fig.1). Both patients with obvious respiratory failure and those at risk of developing severe diseases were hospitalized during the study period in Japan. Patient characteristics are shown in Table 1. No patient took iron supplementation or was diagnosed with hemochromatosis before admission in this cohort. Moreover, no patients had bleeding complications that might affect iron values when they were admitted. Fifty-five patients were in the non-RF group, and 44 and 37 patients had mild and severe RF, respectively. Patients with mild or severe RF had higher age and more comorbidities than those without RF. Moreover, severe RF patients showed high neutrophil counts and CRP levels and low lymphocyte counts.

**Figure 1.**
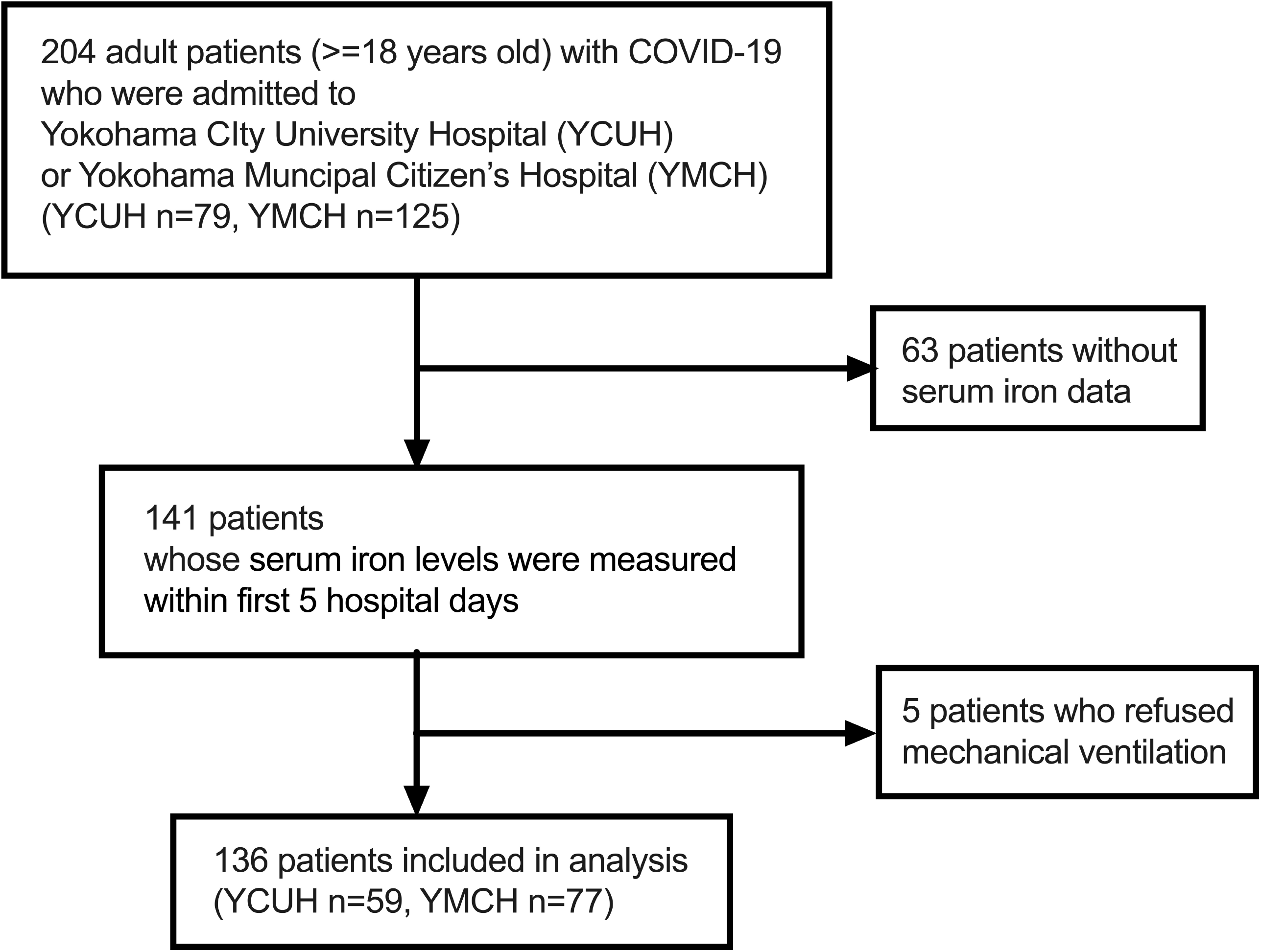
Flow diagram of the patient inclusion.

**Table. 1.** Baseline patient characteristics and commorbidities.

|  | **all** | **non-RF** | **mild RF** | **severe RF** | **p-value** |
| --- | --- | --- | --- | --- | --- |
| Age (years) | 63 (46-73) | 46 (34-59) | 68 (56-75) | 70 (66-78) | *<0.0001 |
| Females/Male | 46/90 | 23/32 | 14/30 | 9/28 | 0.208 |
| Body Mass Index | 22.9 (20.7-25.5) | 22.5 (20.2-25.0) | 22.5 (20.3-25.2) | 23.7 (22.6-26.9) | *0.035 |
| Death (percent [number]) | 5.9 [8] | 0.0 [0] | 0.0 [0] | 21.6 [8] | *<0.0001 |
| **Commorbidities (percent** |  |  |  |  |  |
| **[number])** |  |  |  |  |  |
| Hypertension | 35.3 [48] | 21.8 [12] | 45.5 [20] | 43.2 [16] | *0.008 |
| Diabetes | 27.2 [37] | 12.7 [7] | 36.4 [16] | 37.8 [14] | *0.008 |
| Chronic Respiratory Diseases | 11.0 [15] | 9.1 [5] | 9.0 [4] | 16.2 [6] | 0.4982 |
| Cardiac Diseases | 11.0 [15] | 9.1 [5] | 13.6 [6] | 10.8 [4] | 0.772 |
| Hepatic Diseases | 6.6 [9] | 7.3 [4] | 6.8 [3] | 5.4 [2] | 0.938 |
| Renal Diseases with Hemodialysis | 11 [15] | 7.3 [4] | 13.6 [6] | 13.5 [5] | 0.515 |

\*p<0.05.

**Table. 2.**
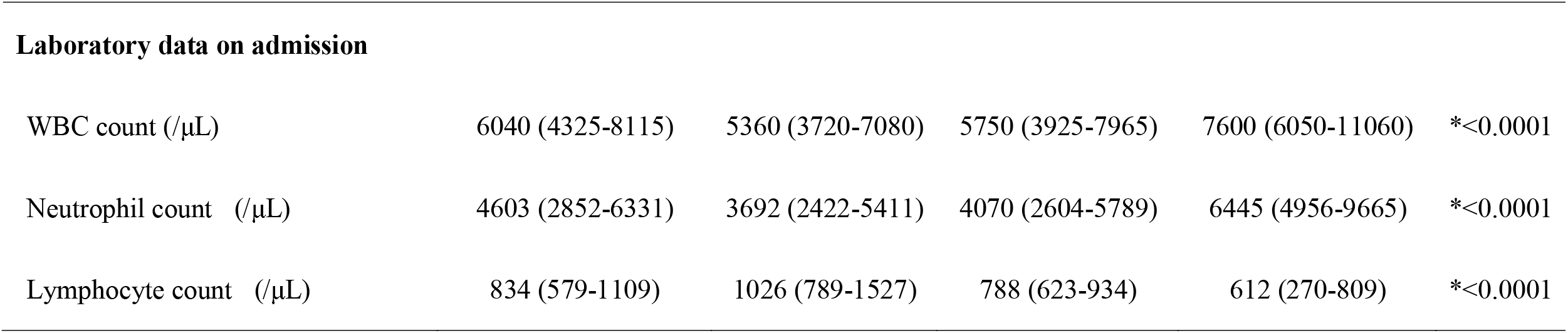
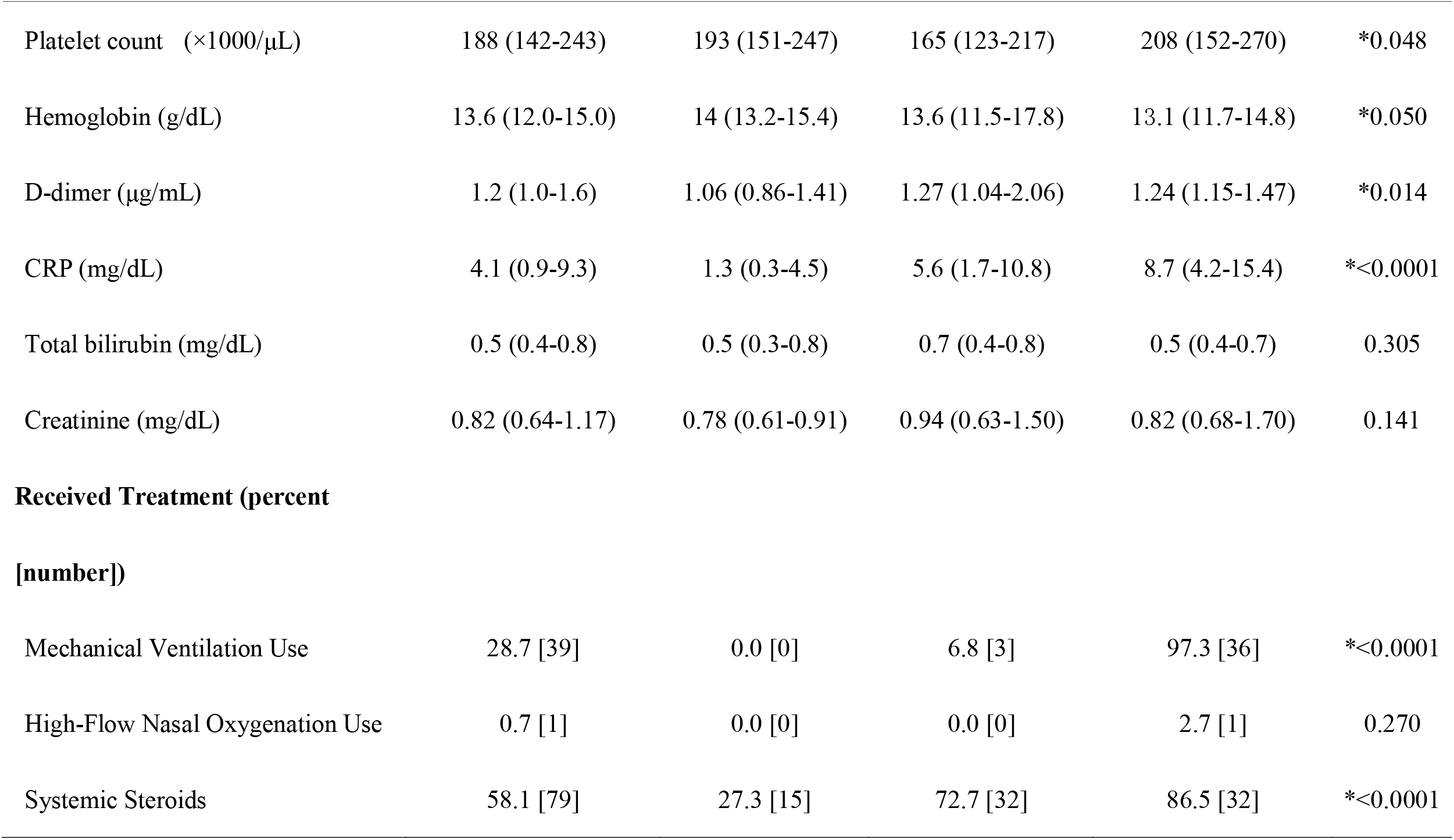
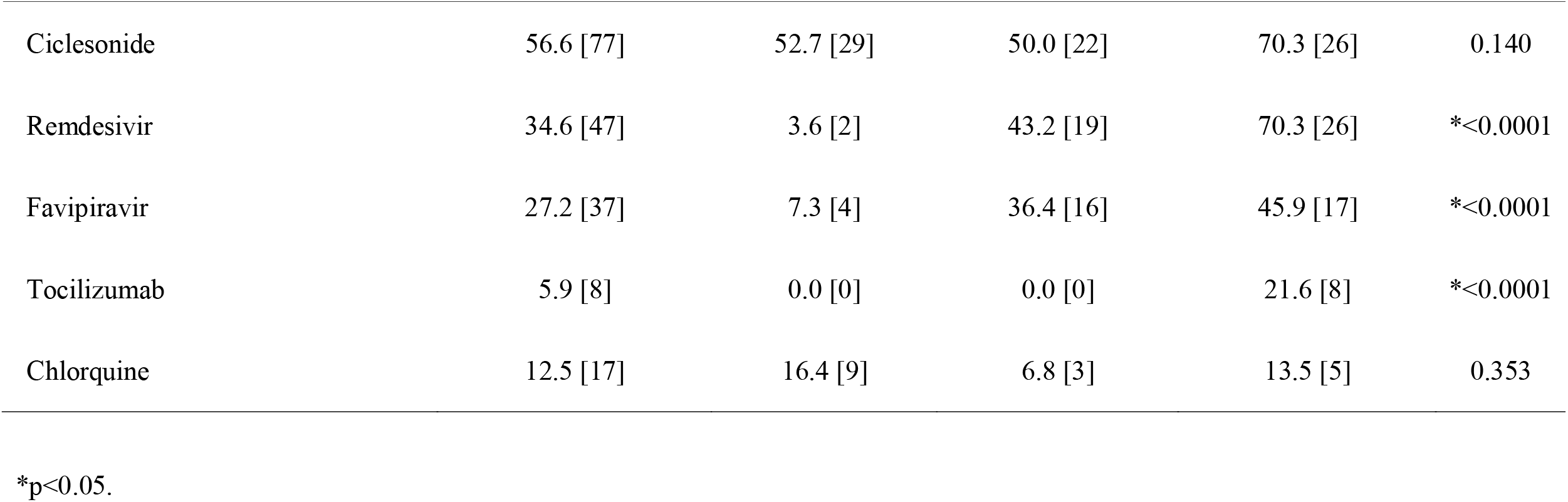
Laboratory data on admission and received treatment during hospiralization.

| **Laboratory data on admission** |  |  |  |  |  |
| --- | --- | --- | --- | --- | --- |
| WBC count (/μL) | 6040 (4325-8115) | 5360 (3720-7080) | 5750 (3925-7965) | 7600 (6050-11060) | *<0.0001 |
| Neutrophil count (/μL) | 4603 (2852-6331) | 3692 (2422-5411) | 4070 (2604-5789) | 6445 (4956-9665) | *<0.0001 |
| Lymphocyte count (/μL) | 834 (579-1109) | 1026 (789-1527) | 788 (623-934) | 612 (270-809) | *<0.0001 |
| Platelet count (×1000/μL) | 188 (142-243) | 193 (151-247) | 165 (123-217) | 208 (152-270) | *0.048 |
| Hemoglobin (g/dL) | 13.6 (12.0-15.0) | 14 (13.2-15.4) | 13.6 (11.5-17.8) | 13.1 (11.7-14.8) | *0.050 |
| D-dimer (μg/mL) | 1.2 (1.0-1.6) | 1.06 (0.86-1.41) | 1.27 (1.04-2.06) | 1.24 (1.15-1.47) | *0.014 |
| CRP (mg/dL) | 4.1 (0.9-9.3) | 1.3 (0.3-4.5) | 5.6 (1.7-10.8) | 8.7 (4.2-15.4) | *<0.0001 |
| Total bilirubin (mg/dL) | 0.5 (0.4-0.8) | 0.5 (0.3-0.8) | 0.7 (0.4-0.8) | 0.5 (0.4-0.7) | 0.305 |
| Creatinine (mg/dL) | 0.82 (0.64-1.17) | 0.78 (0.61-0.91) | 0.94 (0.63-1.50) | 0.82 (0.68-1.70) | 0.141 |
| **Received Treatment (percent**<br>**[number])** |  |  |  |  |  |
| Mechanical Ventilation Use | 28.7 [39] | 0.0 [0] | 6.8 [3] | 97.3 [36] | *<0.0001 |
| High-Flow Nasal Oxygenation Use | 0.7 [1] | 0.0 [0] | 0.0 [0] | 2.7 [1] | 0.270 |
| Systemic Steroids | 58.1 [79] | 27.3 [15] | 72.7 [32] | 86.5 [32] | *<0.0001 |
| Ciclesonide | 56.6 [77] | 52.7 [29] | 50.0 [22] | 70.3 [26] | 0.140 |
| Remdesivir | 34.6 [47] | 3.6 [2] | 43.2 [19] | 70.3 [26] | *<0.0001 |
| Favipiravir | 27.2 [37] | 7.3 [4] | 36.4 [16] | 45.9 [17] | *<0.0001 |
| Tocilizumab | 5.9 [8] | 0.0 [0] | 0.0 [0] | 21.6 [8] | *<0.0001 |
| Chlorquine | 12.5 [17] | 16.4 [9] | 6.8 [3] | 13.5 [5] | 0.353 |

\*p<0.05.

### Iron metabolism in hospitalized COVID-19 patients

Serum iron and ferritin concentrations were measured in all patients within the first 5 days of hospitalization. However, TIBC and TSAT data were missing for 16 patients (4 with mild-moderate RF and 12 with severe RF).

Serum iron levels were significantly lower in the mild RF group (median serum iron level: 24 [interquartile range: 19–42] mg/dL) than in the non-RF (40 [24–80] mg/dL, p=0.019) or severe RF (60 [23.5–87] mg/dL, p=0.009) groups (Fig.2A). No significant difference in iron levels was observed between the severe RF and non-RF groups (p>0.999) (Fig.2A). The TIBC levels significantly decreased with increasing severity (Fig.2B). Consequently, TSAT was significantly higher in patients with severe RF (32.7 [13.9–47.6] %) than in those with non-RF (14.0 [8.4–24.9] %, p=0.012) and mild RF (11.8 [7.8–22.2] %, p<0.001) (Fig.2C). Ferritin levels were significantly increased in patients with RF than in those without RF (Fig.2D), probably due to inflammation, irrespective of iron metabolism. It is well known that iron metabolism markedly varies according to sex. In this study cohort, serum iron levels and TSAT were significantly lower in female patients than in male patients (Supplementary Fig. 1A, B). Moreover, significantly higher TIBC and lower ferritn levels were observed in female patients than in male patients. (Supplementary Fig. 1C, D). Neverttheless, a similar tendency of association between serum iron levels or TSAT and disease severity was observed in both male and female patients (Supplementary Fig. 1E–H).

**Figure. 2.**
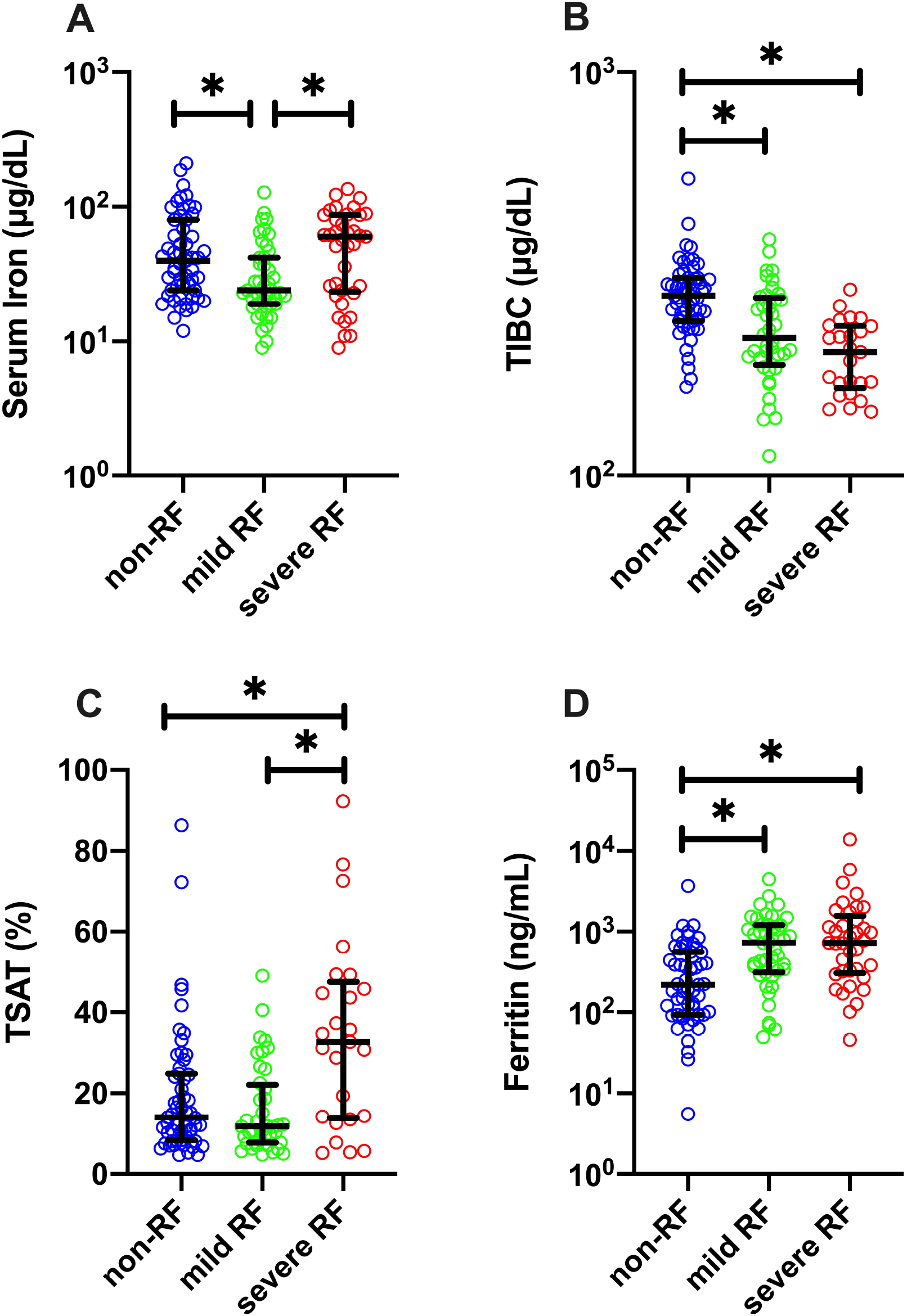
Iron metabolism indicators in hospitalized patients with COVID-19. (A) serum iron, (B) total iron-binding capacity (TIBC), (C) transferrin saturation (TSAT), and (D) ferritin levels. All the values were measured within the first 5 days of hospitalization. Data are presented as median ± interquartile range (IQR). *p<0.05.

Single logistic regression analysis including only patients with RF also demonstrated that higher serum iron (odds ratio 1.93 [95% CI: 1.24–3.15] per 2 folds increase in serum iron level, p=0.005) or TSAT (odds ratio 1.97 [95% CI: 1.38–3.04] per 10 % increase in TSAT, p<0.001) levels were associated with the development of severe RF. Finally, we performed multivariate analysis including only patients with RF, adjusting for age and sex because it is known that the serum iron level is affected by these parameters. Both higher serum iron (odds ratio 2.02 [95% CI: 1.26–3.36] per 2 folds increase in serum iron level, p=0.005) and TSAT (odds ratio 2.00 [95% CI: 1.38–3.16] per 10 % increase in TSAT, p=0.001) levels were independently associated with severe RF among patients with RF.

Finally, we compared serum iron levels between patients with RF who survived and those who did not (Supplementary Fig. 2). Serum iron levels were not significantly different between both groups of patients (28 [19–65] vs. 43 [22–62] mg/dL, p=0.7827).

## Discussion

In the present study, we demonstrated that serum iron levels were significantly lower in COVID-19 patients with mild RF than in those without RF. However, there were no significant differences in iron levels between the non-RF and severe RF groups; that is, we observed a U-shaped association between serum iron levels and disease severity. Additionally, TSAT, which is a potential indicator of catalytic iron level, was significantly higher in patients with severe RF than in other patients. Iron-lowering response upon viral infection is thought to be one of the self-protective host responses^3,12^. Therefore, our results indicate that inadequate iron-lowering response in the severe RF group might have contributed to COVID-19 exacerbation.

Previous studies have demonstrated that serum iron levels are decreased in patients with severe COVID-19^8–11^. In fact, COVID-19 patients with RF (including both mild and severe RF) in this study tended to have lower serum iron levels compared to those without RF. However, in evaluating only patients with RF, higher serum iron levels and TSAT were associated with the development of severe RF. Our study has some strengths compared to other previous studies. First, we analyzed the association between the worst respiratory status and serum iron levels, whereas some previous studies evaluated disease severity on hospital admission. The severity on admission might not necessarily reflect actual disease severity throughout the disease duration. Second, we defined patient outcomes based on a reliable objective criterion, the P/F ratio measured with MV or HFNO. Therefore, we believe that our analysis reflects the actual association between serum iron metabolism and COVID-19 severity.

Serum iron levels are also associated with hypoxic pulmonary vasoconstriction (HPV). It has been reported that iron overload impairs HPV, while chelating serum iron augments HPV^13^. Several reports have shown that ARDS caused by SARS-CoV-2 infection is characterized by severe arterial oxygenation impairment with relatively high lung compliance^14–16^. Therefore, it is assumed that inappropriate pulmonary vascular responses to hypoxia might be involved in the pathophysiology of COVID-19 ARDS^14^. Our observation of higher iron levels in severe COVID-19 cases might be associated with the impairment of HPV and resultant hypoxemia. This suggests that lowering serum iron levels might potentially improve arterial oxygenation in COVID-19-induced severe RF.

Iron metabolism is markedly affected by sex. As expected, our data demonstrated that female patients had lower iron and TSAT levels than male patients. On the other hand, it has been reported that the male sex is a risk for COVID-19 disease severity. Therefore, low iron levels in female patients might be one of the underlying mechanisms of good outcomes. Further studies investigating the association among iron status, sex, and COVID-19 disease severity are warranted.

There were no significant differences in iron levels between patients who survived and those who did not in the present study. The possible reason for this is that mortality was affected by several factors other than iron levels. Moreover, the statistical power is insufficient due to the small number of patients who did not survive (8 of 81 patients with respiratory failure). Further studies including larger cohorts are warranted.

Serum iron levels can be lowered by the administration of iron chelators such as deferoxamine^17^. These iron chelators are already in use to treat iron overload, and drug safety has been established. It is noteworthy that we found three trials evaluating the efficacy of deferoxamine for COVID-19 registered in the clinical trial registration database as of January 2021. In particular, one study was planned to evaluate the preventive effect of deferoxamine on ARDS development. Our data support the rationale of these trials.

This study has some limitations. First, although we performed multivariate analysis, we could only include age and sex as covariates because the number of severe patients in the study cohort was limited. There might be other confounders in the association between serum iron levels and disease severity, although high serum iron levels were still significantly associated with disease severity in the multivariate logistic model, which included several additional covariates (Supplementary Table 1). Second, we could not obtain serum iron levels before SARS-COV-2 infection, although there were no patients with obvious iron metabolism disorders. The pre-infection iron status may likely affect serum iron levels on admission and, consequently, disease severity. Third, we could not obtain the serum concentrations of hepcidin^18^, which is the central regulator of serum iron level upon viral infection. Further studies to evaluate the association between hepcidn levels and COVID-19 severity are necessary.

## Conclusion

In conclusion, we observed a U-shaped association between serum iron levels and RF severity in hospitalized patients with COVID-19. Higher serum iron levels are associated with the development of severe RF in the analysis including only patients with RF, indicating that inadequate response to lower serum iron might be an exacerbating factor for COVID-19.

## Data Availability

The data that support the findings of this study are available from the corresponding author, [KT], upon reasonable request.

## List of abbreviations

ARDS: acute respiratory distress syndrome
COVID-19: coronavirus disease 2019
HFNO: high-flow nasal oxygen therapy
IQR: interquartile range
MV: mechanical ventilation
P/F ratio: arterial oxygen partial pressure / fractional inspired oxygen ratio
RF: respiratory failure
SARS-CoV-2: severe acute respiratory syndrome coronavirus 2
TIBC: total iron binding capacity
TSAT: transferrin saturation
YCUH: Yokohama City University Hospital
YMCH: Yokohama Municipal Citizen’s Hospital

## Declarations

### Ethics approval and consent to participate

The study protocol was reviewed and approved by Yokohama City University Certified Institutional Review Board (approval number: B200700099) and Yokohama Municipal Citizen’s Hospital Institutional Review Board (approval number: 20-09-04). The need for informed consent was waived by Yokohama City University Certified Institutional Review Board and Yokohama Municipal Citizen’s Hospital Institutional Review Board because of the observational design of the study.

### Consent for publication

Not applicable

### Availability of data and materials

The datasets used and/or analyzed during the current study are available from the corresponding author on reasonable request.

### Authors’ contributions

KT conceived and conducted the study, analyzed the data, and wrote the manuscript. YS conducted the study at YMCH and supervised the data collection. YO, FO, and TH supervised the data collection at YCUH. Y Yoshimura, NM, HH, and Y Yamaguchi supervised the data collection at YMCH. YI performed data cleaning and analyzed the data. IT, NT, and TG supervised the study. All authors revised the manuscript.

## Acknowledgments

The authors wish to thank Ms. Satsuki Kusu at Yokohama City University Center for Novel and Exploratory Clinical Trials for supporting the development of the study protocol. We also thank Dr. Takahiro Mihara at Yokohama City University Graduate School of Data Science for providing expert advice on statistical analysis.

