## Supplemental Figure 1 for "The U-shaped association of serum iron level with disease severity in adult hospitalized patients with COVID-19"

**Supplementary Figure. 1**

Comparisons of (A) serum iron, (B) transferrin saturation (TSAT), (C) total iron binding capacity (TIBC), and (D) ferritin levels between male and female patients.

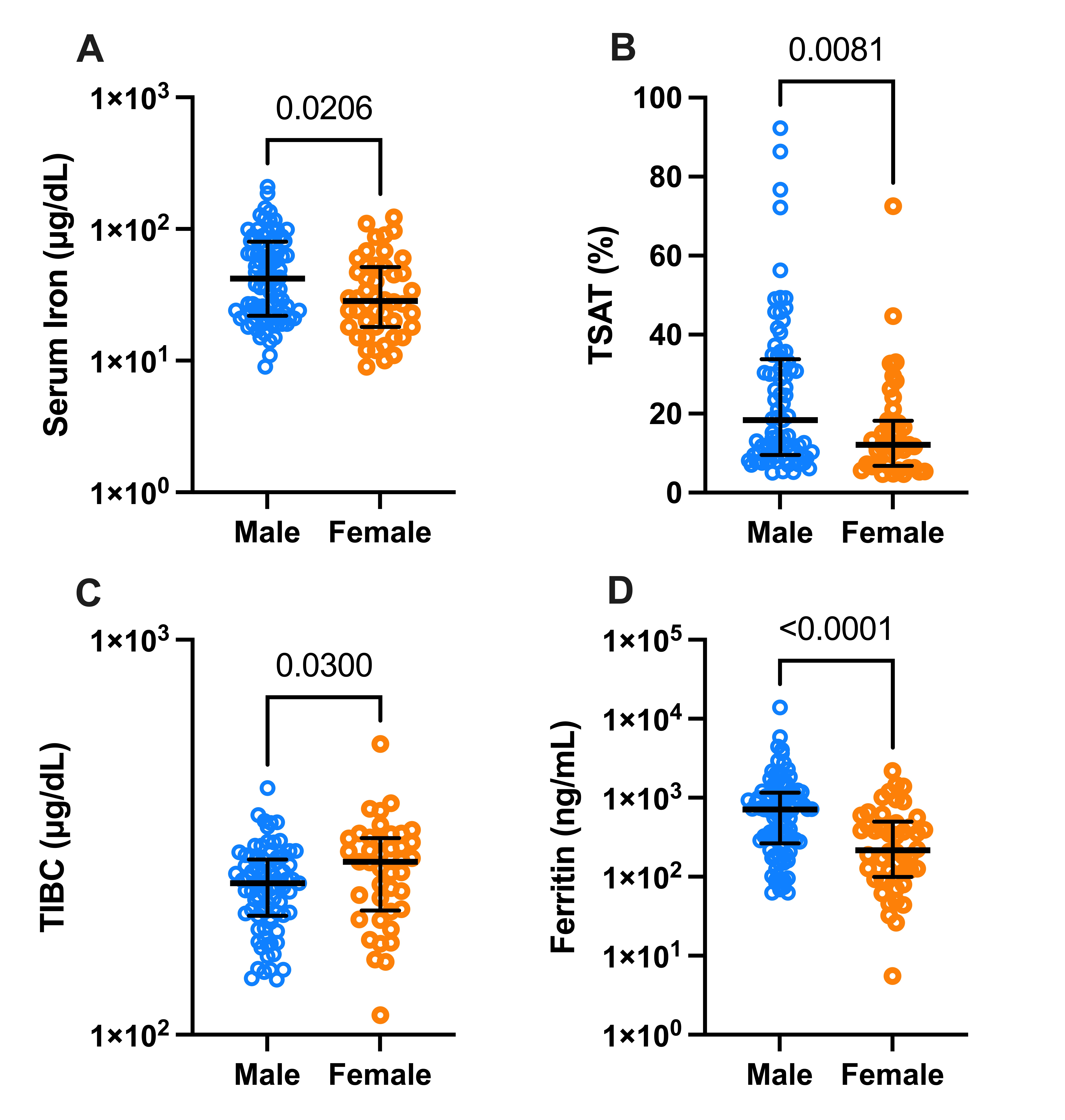

(E) Serum iron and (F) TSAT levels in male patients.

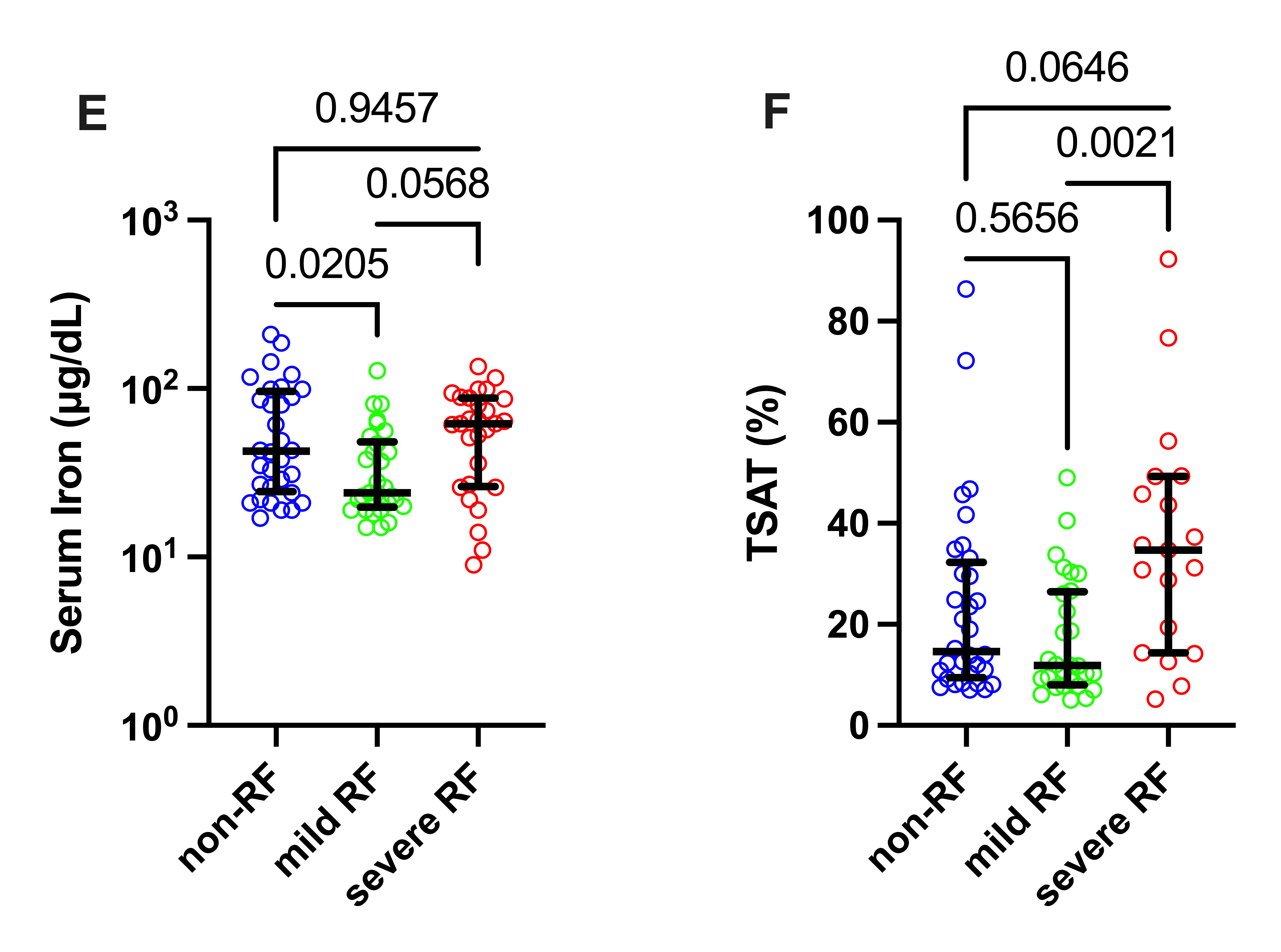

(G) Serum iron and (H) TSAT levels in female patients.

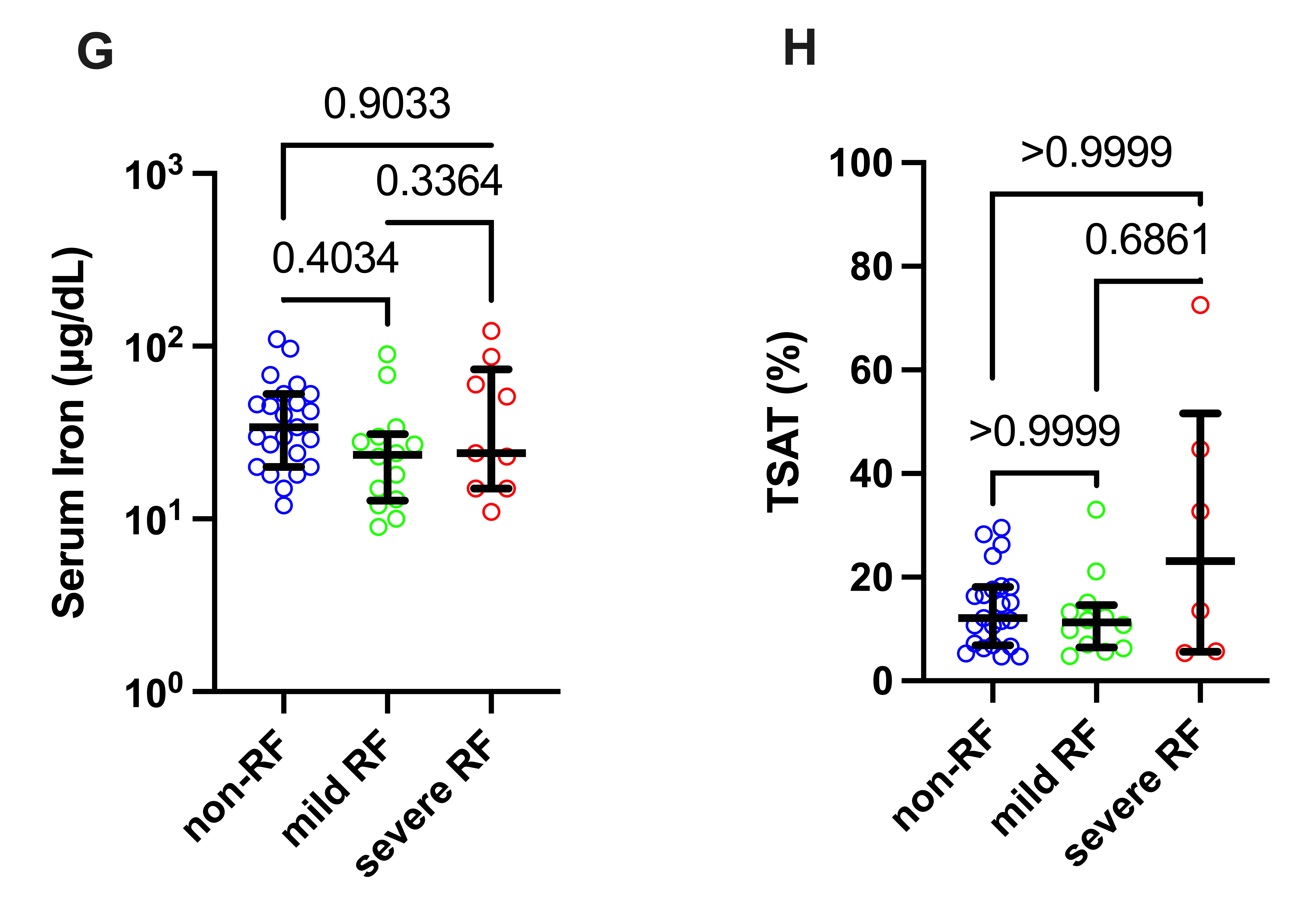
