## Supplemental Figure 2 for "The U-shaped association of serum iron level with disease severity in adult hospitalized patients with COVID-19"

**Supplementary Figure. 2**

Comparisons of serum iron levels between survived and non-survived patients.

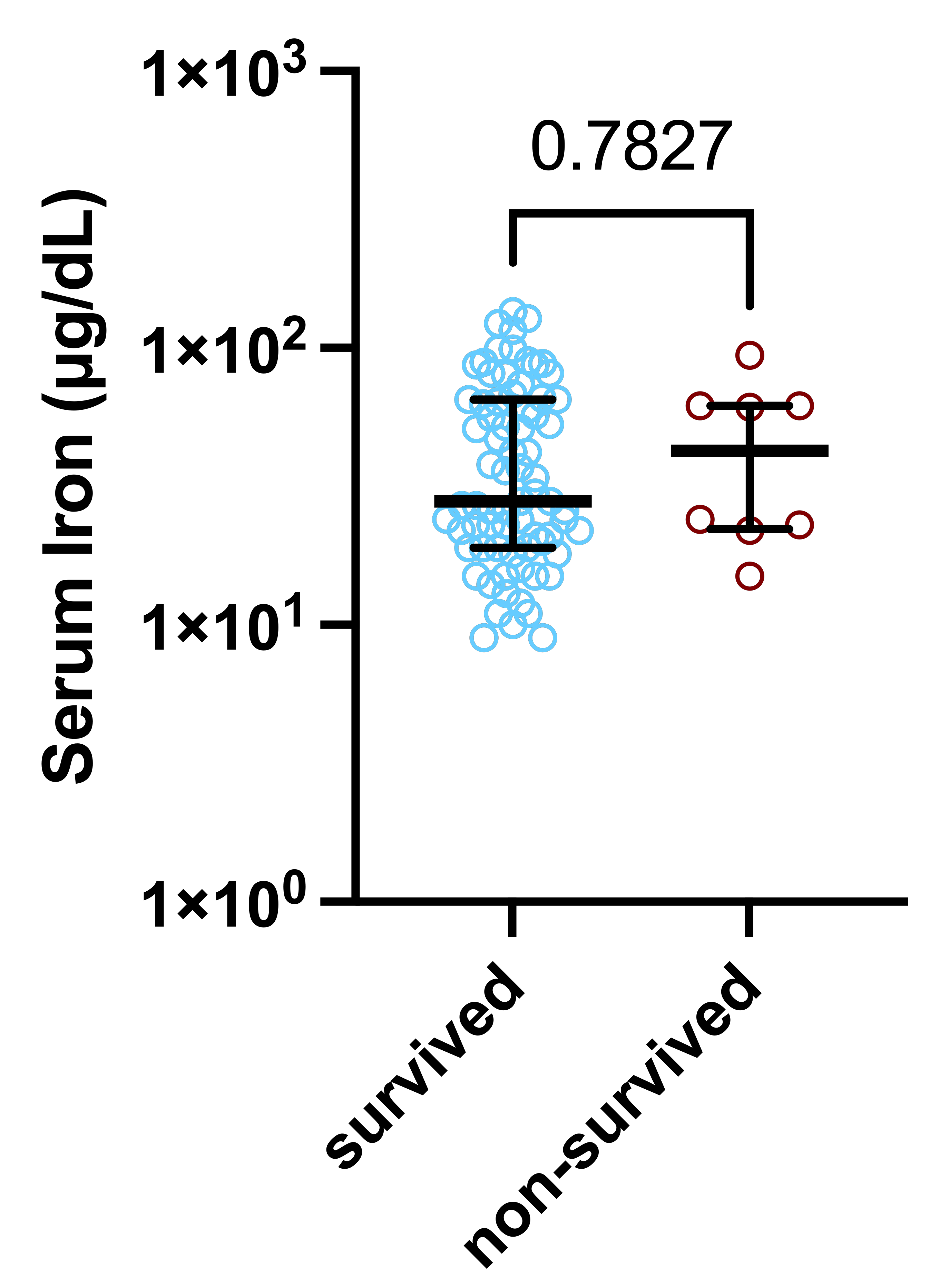
