## Supplemental Table 1 for "The U-shaped association of serum iron level with disease severity in adult hospitalized patients with COVID-19"

**Supplementary Table.1**

Multivariate logistic analysis of risk factors for developing severe disease in hospitalized COVID-19 patients with respiratory failure

| **Variables** | **Odds Ratio (95% CI)** | **P Value** |
| --- | --- | --- |
| Age (per increase of 10 years old) | 1.492 (0.9433 to 2.473) | 0.0994 |
| Female sex | 0.6798 (0.2020 to 2.188) | 0.5207 |
| BMI (per increase of 1) | 1.079 (0.9385 to 1.244) | 0.2857 |
| Hypertension | 0.7849 (0.2765 to 2.200) | 0.6445 |
| Diabetes Mellitus | 1.280 (0.4374 to 3.828) | 0.6520 |
| Chronic Respiratory Diseases | 1.050 (0.2185 to 5.180) | 0.9510 |
| Serum Iron (per 2 folds increase) | 1.845 (1.097 to 3.255) | *0.0258 |

*p<0.05.
